# COVID-19 Vaccination Data Management and Visualization Systems for Improved Decision-Making: Lessons Learnt from Africa CDC Saving Lives and Livelihoods Program

**DOI:** 10.1101/2025.02.14.25322288

**Authors:** Raji Tajudeen, Mosoka Papa Fallah, John Ojo, Tamrat Shaweno, Wondwossen Amanuel, Michael Sileshi, Frehiwot Mulugeta, Moses Bamatura, Dennis Kibiye, Patrick Chanda Kabwe, Senga Sembuche, Ngashi Ngongo, Nebiyu Dereje, Jean Kaseya

**Author notes:** Correspondence: Tamrat Shaweno. Co-first authors (Raji Tajudeen, Mosoka Papa Fallah).

## Abstract

The DHIS2 system enabled real-time tracking of vaccine distribution and administration to facilitate data-driven decisions. Experts from the Africa Centres for Disease Control and Prevention (Africa CDC) Monitoring and Evaluation (M&E) and Management Information System (MIS) teams, with support from the Health Information Systems Program South Africa (HISP-SA), developed the continental COVID-19 vaccination tracking system. Several variables related to COVID-19 vaccination were considered in developing the system. Three-hundred users can access the system at different levels with specific roles and privileges. Four dashboards with high-level summary visualizations were developed for top leadership for decision-making, while pages with detailed programmatic results are available to other users depending on their level of access. Africa CDC staff at different levels with a role-based account can view and interact with the dashboards and make necessary decisions based on the COVID-19 vaccination data from program implementation areas on the continent. The Africa CDC vaccination program dashboard provided essential information for public health officials to monitor the continental COVID-19 vaccination efforts and guide timely decisions. As the impact of COVID-19 is not yet over, the continental tracking of COVID-19 vaccine uptake and dashboard visualizations are used to provide the context of continental COVID-19 vaccination coverage and multiple other metrics that may impact the continental COVID-19 vaccine uptake. The lessons learned during the development and implementation of a continental COVID-19 vaccination tracking and visualization dashboard may be applied across various other public health events of continental and global concern.

**Author Summary:** In our work, we developed a real-time tracking system for COVID-19 vaccination across Africa, using the DHIS2 platform to help guide data-driven decisions. This system, created by experts from the Africa CDC in collaboration with the Health Information Systems Program South Africa (HISP-SA), enables us to monitor and manage COVID-19 vaccine distribution and administration continent-wide. With nearly 300 users from various levels of Africa CDC, the system allows authorized personnel to access relevant vaccination data tailored to their role, making the information easy to interpret and act upon.

Our system includes four interactive dashboards that provide high-level summaries for top leadership and detailed views for program teams. These visual tools empower Africa CDC staff to make informed decisions, track progress, and address challenges in real-time. As COVID-19 remains a global concern, our platform provides crucial insights into vaccine coverage and related health metrics, demonstrating the potential for similar systems to enhance public health responses to future emergencies across Africa and beyond.

## Introduction

The COVID-19 pandemic has highlighted the need for innovative digital health interventions, particularly in vaccines, due to challenges like global inequities, misinformation, and vaccine hesitancy (1). The rapid evolution of public health emergencies (PHEs) demands efficient, real-time solutions to support data-driven decision-making and improve response efforts (2). Digital health technologies have significantly enhanced global health security by enabling timely data collection and analysis, identifying infectious disease trends, and reducing infection risk through remote services (3). Digital tools including data dashboards are being used extensively in the pandemic, collating real-time public health data, including confirmed cases, deaths, and testing figures, to keep the public informed and support policymakers in refining interventions (4). Digital tools have a critical role in increasing the efficiency and effectiveness of the COVID-19 vaccine delivery process and the management of the vaccine program. The speed at which the vaccine is being delivered and administered requires the support of digital technologies that can play a critical role in facilitating the planning, delivery, monitoring, and management of vaccination programs (5). Despite the evident advantages, several challenges hinder the development and implementation of digital solutions in Africa during PHEs. Many African countries face challenges related to internet connectivity, power supply, and access to digital devices, which can impede the widespread adoption of digital health interventions. Ensuring the confidentiality and security of health data remains a significant challenge, particularly with the growing threats of cyberattacks and unauthorized data breaches. Moreover, fragmented health information systems and a lack of standardized protocols hinder seamless data integration across different platforms and institutions (6).

The Saving Lives and Livelihoods (SLL) program, a partnership between the Africa CDC and the Mastercard Foundation, has established information to provide near real-time (NRT) COVID-19 vaccination data for decision-making (7). The Africa CDC and HISP South Africa developed a DHIS2-based system to aid in the distribution and use of COVID-19 vaccines in African countries. Notably, COVID-19 dashboards have also been effectively used in different settings to track COVID-19 cases, deaths, and testing throughout the COVID-19 pandemic (8–14). As the world prepares for future emerging and re-emerging pandemics, there is a need to establish strengthened proactive case identification, efficient triaging protocol, and reporting systems using innovative digital technologies in regions where outbreaks are likely to occur, to increase swift response to future epidemics before they evolve into pandemics (15). Integrating digital health solutions with immunization strategies holds immense potential for improving immunization coverage and monitoring, especially in the post-COVID-19 era (16). Strengthening Africa’s digital health infrastructure can help improve real-time surveillance, enhance cross-border coordination, and ensure equitable access to healthcare interventions. Herein, we describe the overall process and critical lessons learned in the development and implementation of the Africa CDC SLL COVID-19 vaccination dashboard, highlighting its role in improving vaccination monitoring and public health response in Africa while emphasizing the importance of scaling up digital solutions for future public health threats.

## Methods and Materials

### Data sources

COVID-19 vaccination data is sourced from multiple sources, including SLL implementing partners (IPs), Member States, and other public sources such as the weekly COVID-19 logistics data from the World Health Organization (WHO). The DHIS2 platform collects and visualizes this data, providing insights into program implementation and vaccine administration (17). Implementing Partners, contracted under the SLL program, share data validated by the health authority of the implementing Member State. Data entry and submission to Africa CDC are done weekly, monthly, and quarterly, with data reporting on vaccination figures, total number of vaccine doses administered by the SLL program, risk communication and community engagement, and safety surveillance, respectively. Each SLL IP supports programmatic activities in a Member State, and the uploaded data undergoes approval by designated Africa CDC National Coordinators (NCs).

The COVID-19 vaccination dashboard presents detailed information on vaccine administration, partnership activities, health worker training, vaccine safety, RCCE, job creation, and project management-related data. The dashboard visually organizes data according to the user and uses standardized Key Performance Indicators (KPIs) to measure performance against targets. Member States with low performance are the focus of the Daily War Room meetings to identify the root causes and seek alternative strategies for improved performance.

### Contents and process of the dashboard development

We used an open-source web-based software platform called DHIS2 software (18) for the dashboard development. The dashboard was designed in phases, with subject matter experts meeting weekly to draft business specification requirements (BSR). The BSR was then handed over to HISP-SA. The initial draft was approved for internal trial use, data quality assurance checks, and large-area server security checks.

The dashboard explores and summarizes vaccination data and attributes, including vaccination coverage, number of persons vaccinated disaggregated by gender, vaccine types used, and risk communication and community engagement (RCCE) activities for Member States implementing the SLL program. The dashboard is structured around 38 datasets, containing 459 distinct data elements and 137 indicators. The dashboards include metrics such as trained vaccinators, vaccine stocks, vaccines nearing expiry, AEFI data, trained community mobilizers, community leaders engaged in delivering COVID-19 prevention messages, etc. (Table 1). The dashboard uses API connectivity for efficient data sharing.

**Table 1.** Categories and metrics included in SLL COVID-19 Vaccination Dashboard.

| Category | Metrics |
| --- | --- |
| COVID-19 vaccine administration | - Country doses administered weekly<br>- Doses administered monthly<br>- Doses administered by SLL-supported CVCs<br>- Doses administered per population<br>- Doses administered per target population<br>- Partner - Total doses administered by the SLL program<br>- Target of doses administered<br>- Vaccination status by sex<br>- Total number of CVCs<br>- Doses received |
|  | - Doses wasted<br>- Doses in the stock (AstraZeneca, J&J, Moderna, Pfizer/BioNTech, Sinopharm, Sinovac)<br>- Ancillary units received<br>- Target number of doses to be administered |
| RCCE | - Community health workers performing RCCE<br>- Community health workers performing RCCE at CVC site<br>- People reached through RCCE activities via communication channel<br>- People trained on RCCE approaches and the COVID-19 vaccine<br>- RCCE area of support<br>- Partner community health workers trained<br>- Partner vaccinators trained<br>- Number of COVID-19 vaccination mass campaign conducted<br>- RI sites that have integrated COVID-19 vaccination into PHCs<br>- Partner-Number of vaccination teams supporting campaigns |
| Staffing | - Total number of staffs working onsite<br>- Country data managers trained |
|  | - Frontline healthcare workers eligible for COVID-19 vaccination<br>- Jobs created by SLL<br>- People trained by National Societies<br>- Youth employment |
| Statistics | - Fully vaccination rate<br>- Proportion of wastage of PEE during transportation<br>- Share of COVID-19 cases sequenced<br>- Share of the population fully vaccinated<br>- Share of target population fully vaccinated<br>- Number of COVID-19-related technical reports published<br>- Scientific presentations in technical meetings |
| AEFI | - AEFIs reported by SLL partners<br>- Confirmed AEFIs following vaccination<br>- Deaths from AEFIs<br>- Reported or suspected (AEFI and adverse AEFI) following COVID-19 vaccination<br>- Serious AEFI reported by SLL partners<br>- Serious AEFIs investigated within 48 hours |
| Procurement | - Total requests received /monthly/quarterly<br>- Vaccine doses delivered<br>- Doses procured by the SLL program<br>- Country number of doses received |

### Access to online data and validation process

The Africa CDC SLL DHIS2 dashboard allows key stakeholders to access data through authentication and on the Account Creation-UAC form, username name and password shared via email and WhatsApp message, respectively. The dashboard has four-page views for top leadership and a reduced number for other staff. Data entry access is granted to national-level data entry, with no right for other managers or staff. Vaccination data is entered into the system using the “Data Entry App” and validated using the “*Run validation*” command (Fig 1).

**Fig 1.**
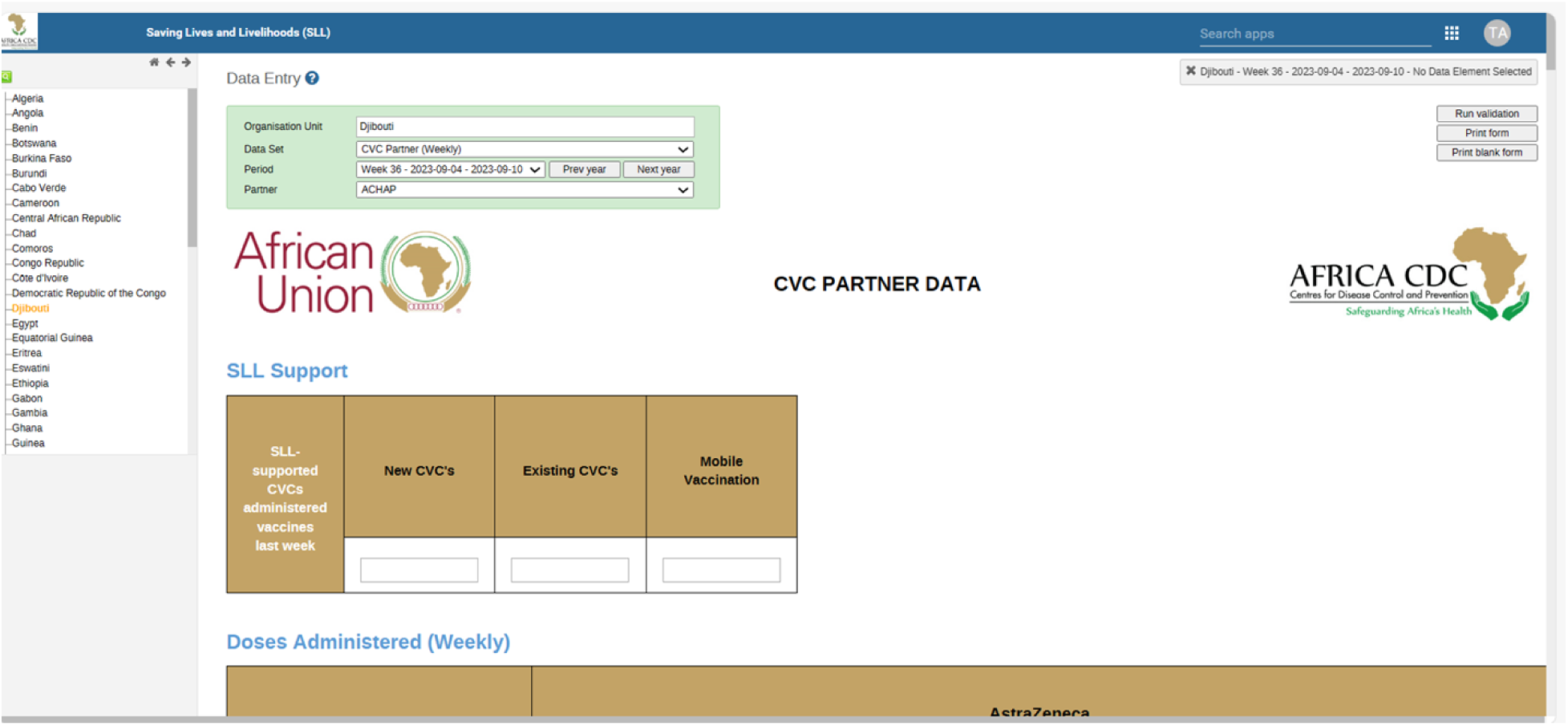
The online Africa CDC COVID-19 vaccination data entry form.

### Timelines for uploading COVID-19 vaccination data into the DHIS2 system

The DHIS2 system manages COVID-10 vaccination data, including weekly COVID-19 Vaccination Centres (CVCs) data, monthly RCCE data, and quarterly pharmacovigilance data. CVC data comes from fixed health facilities and mobile vaccination centres and is reported weekly. National Coordinators review data submissions and either query or approve anomalous data. The system locks in the values to prevent further amendments. NCs analyze partners’ data submissions, troubleshoot, and identify anomalous values. RCCE data is submitted on the 15^th^ of the next Month, and NCs review and lock in values. If anomalous data is found, NCs engage IPs to try to understand the root causes and correct the data., DHIS2 automatically verifies data completeness and computes reporting timeliness based on pre-programmed information.

Data is displayed in dashboards within hours, and if data is not shared within the set timeline, an escalation cascade is initiated. If data inconsistencies are detected, a non-compliance state is triggered. Follow-ups are made via email and notifications are sent to NCs in the respective countries to explore the root cause.

### Data visualization, and decision-making process

The Africa CDC and Mastercard Foundation use the DHIS2 system to track and monitor data inputted into the COVID-19 vaccination program. The system features data entry, approval processes, and data visualization apps.apps shown in [Fig. 2].

**Fig 2.**
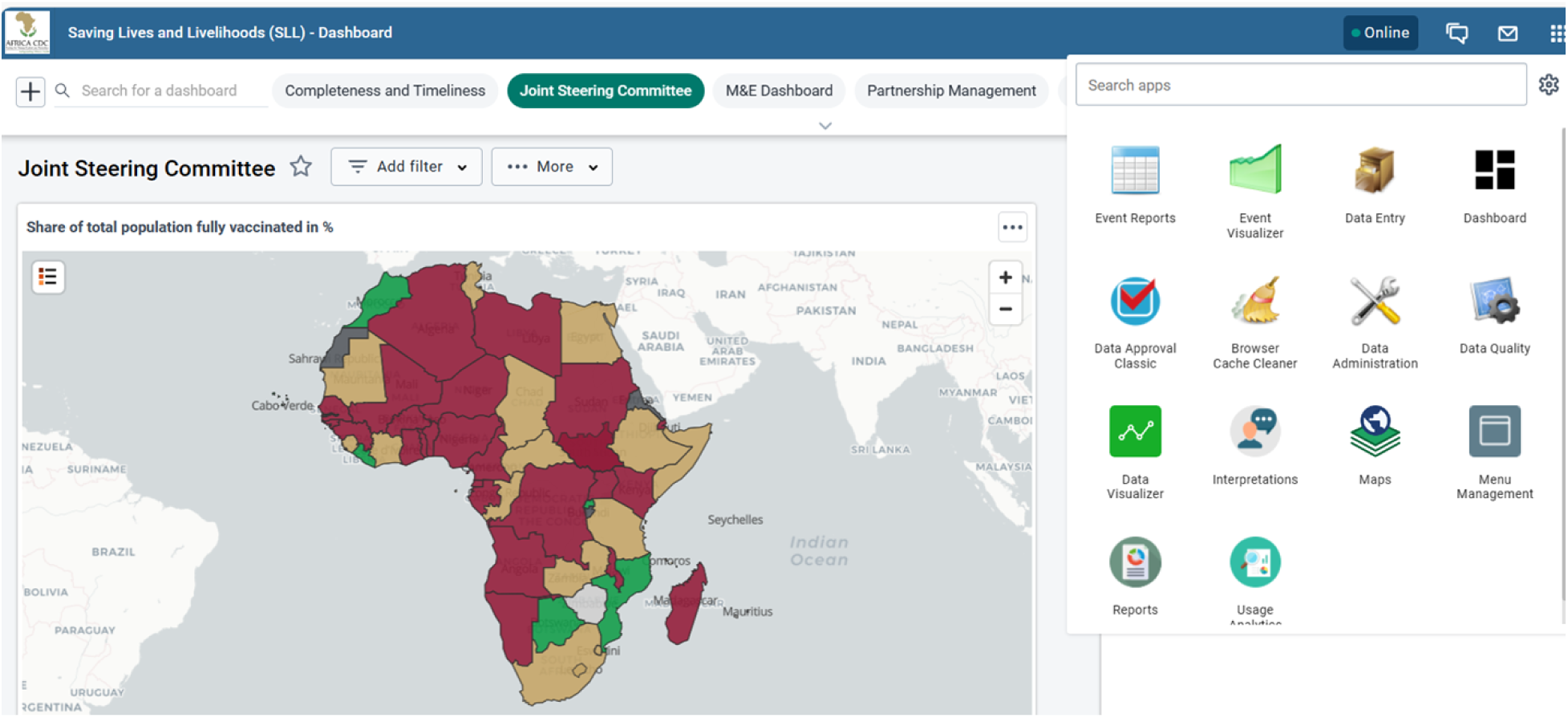
Key features of Africa CDC SLL Program DHIS2 system Apps: data entry, data approval process, and data visualizer apps.

The Data Visualizer app allows users to create various charts directly within DHIS2, with specific layout restrictions and use cases.

The visualization can be viewed as pivot tables, columns, stacked columns, bars, stacked bars, and line graphs, and can be downloaded as images, tables, Excel data, or other types of formats for further analysis. COVID-19 data visualization improves the digitalization and quality of data and enabling better data use and performance monitoring in Member States. It avoids report delay and disintegrated reporting systems, preventing tedious work of aggregation and human error. The enhanced visualization system has successfully rectified discrepancies observed in data between platforms. Different types of charts available in DHIS2 that can be used to visualize COVID-19 data are shown in [Fig. 3].

**Fig 3.**
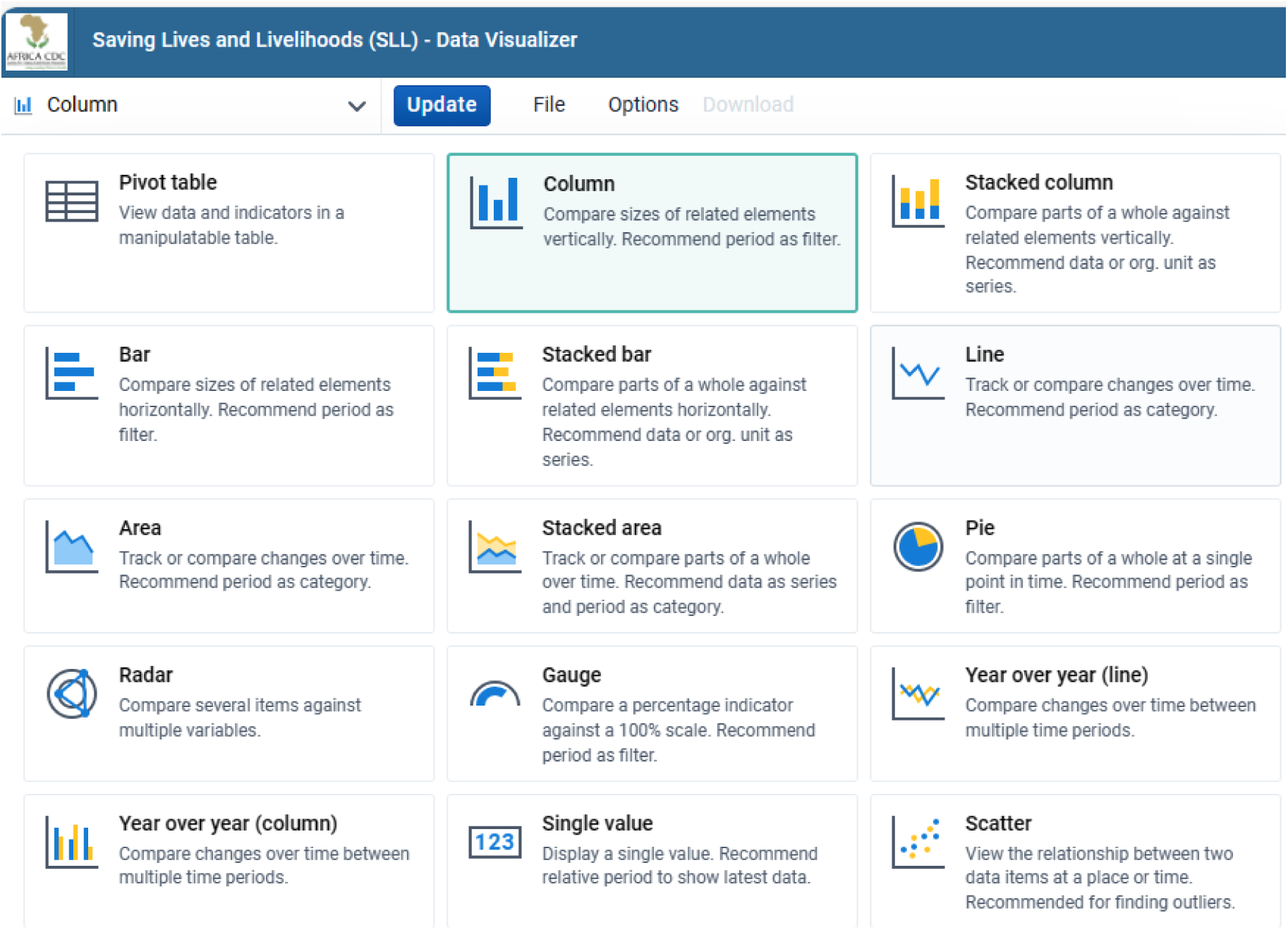
Types of data visualization apps available in DHIS2.

### Ethical consideration

The project on real-time COVID-19 vaccine tracking via the DHIS2 system did not require ethical approval. The initiative involved creating a digital platform for monitoring vaccination efforts across Africa, with data aggregated for public health decision-making by authorized Africa CDC personnel. As no personally identifiable or sensitive individual-level health data were collected, and users operated within defined professional roles, the project did not qualify as human subject research, hence, an ethical waiver was appropriate.

## Result

### The decision-making process

There are four high-level decision-making bodies in the SLL program, each with a dashboard containing critical information for quick decision-making. These dashboards are named after the corresponding decision body and include the Joint Steering Committee Dashboard, the Program Management Unit (PMU) Daily War Room Dashboard, the Vaccine Taskforce Dashboard, and the Partnership Management Dashboard.

### Joint Steering Committee Dashboard

The Joint Steering Committee, led by Africa CDC and the Mastercard Foundation, is the highest-level decision-making entity for the SLL program. It oversees program implementation tracking and uses the Joint Steering Committee dashboard to make timely corrections based on indicators. The dashboard monitors and analyzes overarching SLL and Member State performance against targets such as the share of the total population vaccinated on the continent. An example map shows **[Fig.4]** that eight Member States had achieved 100% full vaccination of their population, 12 achieved 70%, and the remaining countries had below 70%. The dashboard allows the committee to make actionable decisions across the program sites to ensure Member States with poor performance are compared to better-performing ones.

**Fig 4.**
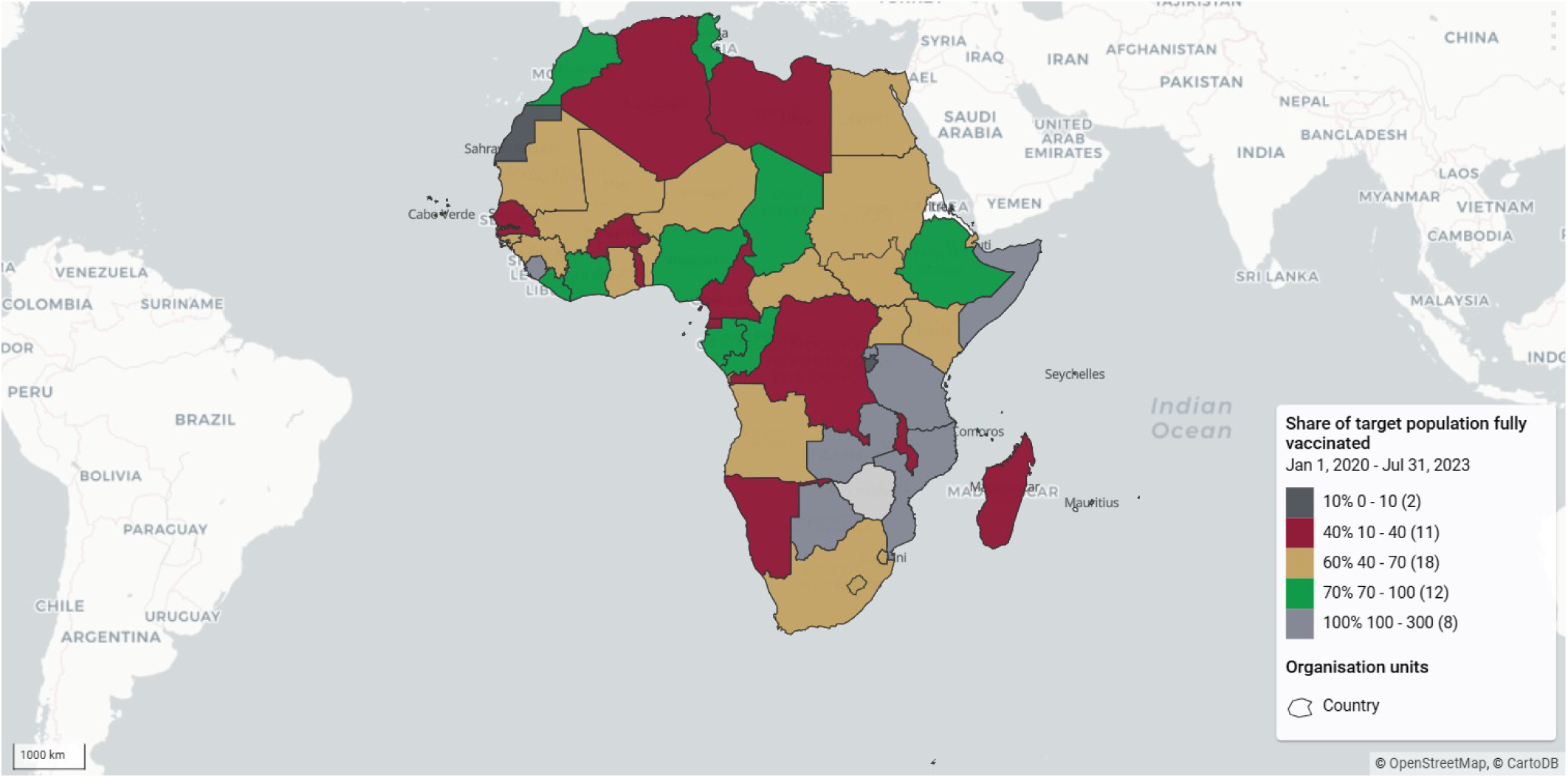
Share of target population fully vaccinated in percentage.

The Joint Steering Committee can use the vaccination dashboards disaggregated by region. The below outputs show the cumulative performance against targets in one of the five African Union regional coordinating centers, the Eastern AU region. From the dashboard in **[Fig.5]**, the overall regional performance against the target as well as individual country performance against the target can be easily tracked, and timely decisions can be made.

**Fig 5.**
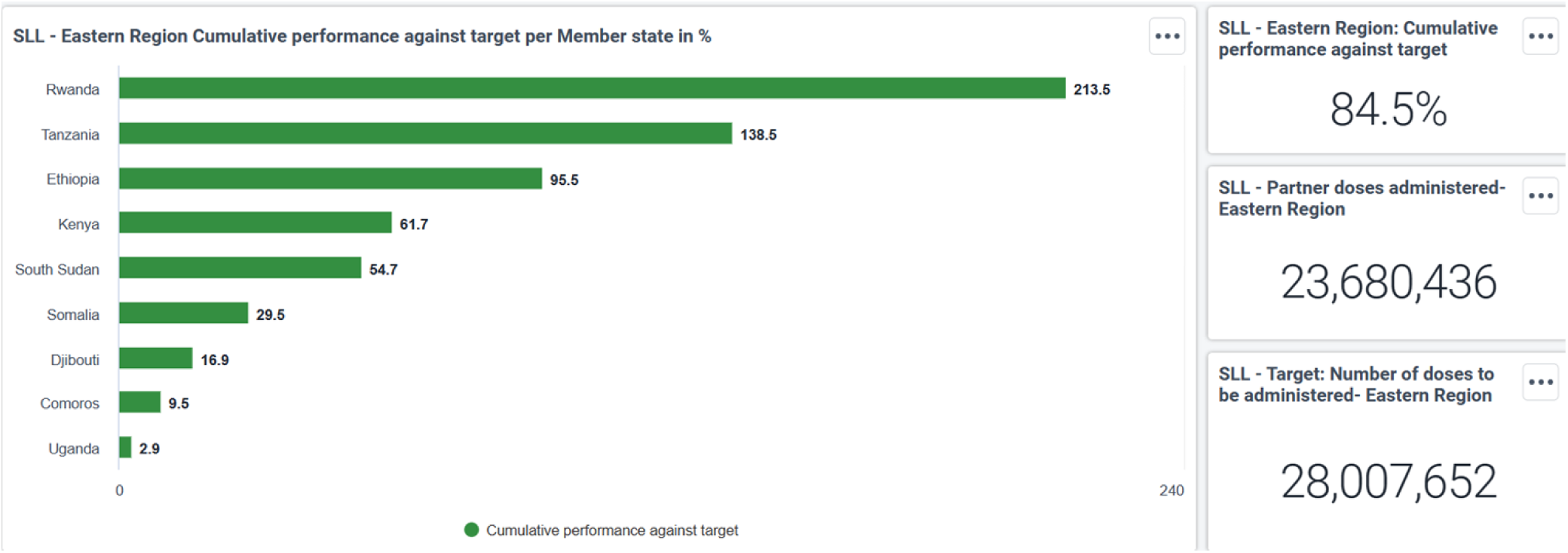
SLL - Eastern Region Cumulative performance against target per Member state in %.

The Joint Steering Committee dashboard displays a chart indicating the Saving Lives and Livelihoods program’s contribution to COVID-19 vaccination administration in each Member State. From the chart, SLL contributed 31.2% of vaccines administered in Rwanda, followed by Ethiopia (26%).

### Program Management Unit (PMU) Daily War Room

The PMU is responsible for coordinating the SLL program, monitoring performance, and reporting to the governing bodies. It holds meetings to develop plans, manage relationships with the Mastercard Foundation, support teams, and serve as the single source of truth. The PMU Daily War Room dashboard assesses regional performance tracking performance against targets, and displays weekly target doses to be administered vs Partner doses administered in each implementing Member State. It also shows vaccine administration per existing and mobile CVCs for the last 12 weeks.

### Vaccine Taskforce dashboard

The main objective of the Vaccine Taskforce dashboard is to monitor the overall continental and implementing Member States’ performance against targets to enable course correction and make data-driven decisions.

### Partnership Management dashboard

The Partnership Management team is responsible for setting up partners’ agreements and supporting partners to clarify contract-related queries during implementation. The team performs activities to include selecting partners and setting up partners’ agreements, supporting partners to define their high-level scope of work and interaction model, monitoring compliance-related or contractual issues encountered by partners during implementation, and overseeing sourcing and procurement by partners. The main objective of the Partnership Management dashboard is to monitor partner performance across each Member State.

### Monitoring and evaluation dashboard

Effective performance monitoring and evaluation is done by collaborating with all parties involved and viewing data collection and validation as a joint responsibility. Africa CDC’s monitoring and evaluation team oversees and ensures each stakeholder plays a role in collecting, reporting, and validating data and activity pictures. Moreover, the M&E dashboard tracked the weekly COVID-19 vaccine doses administered in each Member State.

## Discussion

The rapid onset of the COVID-19 pandemic necessitated that countries worldwide quickly implement data-reporting systems, many of which relied on digital tools. This urgency, alongside the strain on health systems, led to increased collaboration between innovators and governments to adapt digital technologies for various functions, including case management, contact tracing, evidence-based surveillance, risk communication, and vaccine distribution (19). This study presents the COVID-19 vaccination data management and visualization systems that enhanced decision-making at the continental level in Africa. Key insights from the Africa CDC’s Saving Lives and Livelihoods (SLL) Program offer valuable lessons for the global community, demonstrating the power of digital solutions in public health management.

### The SLL Program’s Decision-Making Framework

A significant innovation of the SLL program was establishing four primary decision-making bodies supported by specialized dashboards to facilitate data-driven action. This approach aligns with the broader trend in public health of using data visualization tools to enhance decision-making processes. Research has shown that real-time dashboards are essential for managing health interventions during crises like the COVID-19 pandemic (20) and are crucial for transparency and stakeholder engagement (21). Thus, the SLL program reflects a global shift towards data-centric governance, leveraging technology to drive effective program outcomes.

At the apex of the SLL program’s governance is the Joint Steering Committee, led by Africa CDC and the Mastercard Foundation, focusing on monitoring and adjusting program implementation. This structure is pivotal in meeting vaccination targets, a goal often highlighted in public health studies. For instance, research suggests that effective leadership and monitoring bodies can significantly enhance vaccination coverage (22). The program’s dashboard, with regional breakdowns showcasing performance variations among countries, resonates with findings that disparities in vaccination uptake often stem from differing health system capacities and regional policies (23). This evidence underscores the need for tailored support to improve vaccination rates across all Member States.

The Program Management Unit (PMU) functions as the operational core of the SLL program, ensuring coordination and progress reporting. The PMU’s Daily War Room Dashboard, which tracks vaccination targets and doses administered weekly, aligns with best practices in health intervention management. Continuous performance tracking allows timely adjustments, enhancing overall program outcomes (24). Additionally, the emphasis on utilizing both permanent and mobile clinics for vaccine administration highlights the importance of accessibility in vaccination campaigns, particularly in underserved regions (25), demonstrating a commitment to comprehensive coverage.

The Vaccine Taskforce Dashboard provides a comprehensive view of vaccination efforts across Member States, enabling data-driven adjustments where needed. This adaptability is supported by studies that emphasize the necessity of flexible frameworks in public health (26), which allow for rapid responses to challenges. The “bird’s-eye view” approach of the Taskforce Dashboard facilitates the identification of trends and coverage gaps, fostering proactive decision-making to achieve program objectives. This perspective is supported by research indicating that comprehensive data collection enhances intervention effectiveness (27).

The Partnership Management Dashboard plays a critical role in monitoring partner performance across Member States, ensuring compliance and effective collaboration. Successful health interventions often rely on strong partner relationships and clear communication, with studies showing that effective partnership management correlates with improved program outcomes (28). The dashboard’s emphasis on tracking compliance and supporting agreements highlights the role of robust contractual frameworks in fostering partner accountability (29), which is essential for optimizing health service delivery.

The Monitoring and Evaluation (M&E) component of the SLL program, led by Africa CDC, focuses on collaborative data collection and validation, a trend increasingly advocated in public health to improve data reliability. Inclusive M&E strategies yield more comprehensive insights into program effectiveness (30). The program’s weekly tracking of vaccine administration to maintain accountability and transparency echoes best practices in health monitoring, highlighting the importance of timely reporting for stakeholder trust (33). This collaborative approach enhances the program’s adaptability, strengthening its response to evolving vaccination challenges.

### Lessons learned

The SLL program’s strategic use of decision-making bodies and dashboards reflects a commitment to effective program management, accountability, and data-driven decision-making. The COVID-19 vaccination dashboard, developed by Africa CDC with support from HISP South Africa, has become an essential tool for real-time monitoring of vaccine procurement, administration, and data management, informing Africa CDC and Mastercard leadership on vaccination progress across the continent. The dashboard’s interactive features allow for rapid filtering and visualization of data, supporting decision-makers in implementing COVID-19 vaccination strategies at all levels. On average, since June 2022, SLL program implementers have administered approximately 582,000 doses weekly to eligible populations across Member States.

Near real-time data capturing and digital governance have accelerated data sharing, analysis, and utilization, enabling implementers and Member States to monitor vaccination progress, identify gaps, and manage vaccine supply and logistics. This capacity for real-time adjustments has proven vital for vaccine equity and accessibility, essential components of a successful continental vaccination strategy. The COVID-19 vaccination dashboard continues to evolve based on the SLL vaccination protocols, supporting targeted vaccination campaigns and fostering equitable vaccine uptake across Member States. These insights suggest that adaptable, interactive dashboards have the potential to transform the response to public health challenges, enabling effective distribution and uptake of vaccinations and enhancing readiness for future disease outbreaks.

### Challenges and Limitations

Despite successes, several challenges were encountered. Limited data visualization literacy among some public health professionals necessitated training to improve data use and interpretation skills. Data interoperability presented another challenge, as inconsistent data formats across systems hampered integration. Other limitations included incomplete vaccine data in remote areas, internet connectivity issues, and the lack of dedicated data managers in some Member States, all of which impacted the timeliness of data entry and decision-making.

### Conclusion

The SLL program demonstrates the importance of strategic decision-making frameworks and digital dashboards in supporting program management and fostering a data-driven, adaptable approach to public health. By examining these strategies in light of current public health literature, the program emerges as an exemplary model within the paradigm of adaptability, collaboration, and transparency in achieving health objectives. Rapid, multi-level communication between managers, staff, and implementing partners has enabled the Africa CDC to improve COVID-19 vaccine uptake, manage ongoing challenges, and prioritize Member States’ needs. Translating this framework of digital immunization dashboards to other public health areas could further promote health equity and advance digital health initiatives across the continent. Moreover, Integrating digital health solutions with immunization strategies can enhance coverage and monitoring, improving public health outcomes. In the post-COVID-19 era, leveraging digital innovations can make immunization programs more efficient and effective, promoting better health for all.

## Acknowledgments

We acknowledge Africa CDC’s leadership and expertise in various workstreams, for developing and making use of the COVID-19 vaccination tracking dashboard to ease the decision-making at various structures of the continental health agency. We are grateful to the Mastercard Foundation for funding the initiative and HISP South Africa (HISP-SA) for the invaluable technical support for the development of the continental COVID-19 vaccination tracking system. Moreover, we thank the Implementing Partners and Member States for their dedication to submitting COVID-19 data with full quality and integrity.

## Data Availability Statement

All data are in the manuscript files.

## Author Contribution

**Conceptualization:** Raji Tajudeen, Mosoka Papa Fallah, Alian Ngashi Ngongo, John Ojo,

**Data curation:** John Ojo, Wondwossen Amanuel, Michael Sileshi, Frehiwot Mulugeta, Moses Bamatura, Dennis Kibiye

**Formal analysis:** Tamrat Shaweno, Nebiyu Dereje

**Funding acquisition:** Raji Tajudeen, Mosoka Papa Fallah, Alian Ngashi Ngongo

**Investigation:** John Ojo, Tamrat Shaweno, Nebiyu Dereje, Wondwossen Amanuel, Michael Sileshi, Frehiwot Mulugeta, Moses Bamatura, Dennis Kibiye, Patrick Chanda Kabwe, Senga Sembuche

**Methodology:** John Ojo, Tamrat Shaweno, Nebiyu Dereje, Wondwossen Amanuel, Michael Sileshi, Frehiwot Mulugeta, Moses Bamatura, Dennis Kibiye

**Project administration:** Raji Tajudeen, Mosoka Papa Fallah, John Ojo

**Software:** Tamrat Shaweno, Nebiyu Dereje, John Ojo, Wondwossen Amanuel, Michael Sileshi, Frehiwot Mulugeta

**Supervision:** Nebiyu Dereje, Jean Kaseya

**Validation:** Mosoka Papa Fallah, John Ojo, Patrick Chanda Kabwe, Senga Sembuche

**Visualization:** John Ojo, Tamrat Shaweno, Nebiyu Dereje, Wondwossen Amanuel, Michael Sileshi, Frehiwot Mulugeta, Moses Bamatura, Dennis Kibiye

**Writing – Original Draft Preparation:** Tamrat Shaweno, Nebiyu Dereje, John Ojo, Wondwossen Amanuel, Michael Sileshi, Frehiwot Mulugeta, Moses Bamatura, Dennis Kibiye, Patrick Chanda Kabwe

**Writing – Review & Editing:** Mosoka Papa Fallah, John Ojo, Tamrat Shaweno, Nebiyu Dereje, Jean Kaseya

## Abbreviation List

Africa CDC: Africa Centres for Disease Control and Prevention
AEFIs: Adverse Effects Following Immunizations
BSR: business specification requirements
CSV: Comma Separated Values
HTML: Hypertext Markup Language
IP: Implementing Partner
JSON: JavaScript Object Notation
KPIs: Key Performance Indicators
M&E: Monitoring and Evaluation
NCs: National Coordinators
SLL: Saving Lives and Livelihoods
COVAX: COVID-19 Vaccines Global Access
WHO: World Health Organization

